# A novel methodology for the synchronous collection and multimodal visualisation of continuous neurocardiovascular and neuromuscular physiological data in adults with long COVID

**DOI:** 10.1101/2021.12.24.21268370

**Authors:** Feng Xue, Ann Monaghan, Glenn Jennings, Lisa Byrne, Tim Foran, Eoin Duggan, Roman Romero-Ortuno

**Author notes:** Corresponding author: Feng Xue, TROPIC/FRAILMatics office, 2^nd^ Floor, Mercer’s Institute for Successful Ageing (MISA), St James’s Hospital, Dublin 8, Ireland.

## Abstract

Reports suggest that adults with post-COVID-19 syndrome or long COVID may be affected by orthostatic intolerance syndromes, with autonomic nervous system dysfunction as a possible causal factor of neurocardiovascular instability (NCVI). Long COVID can also manifest as prolonged fatigue, which may be linked to neuromuscular function impairment (NMFI). The current clinical assessment for NCVI monitors neurocardiovascular performance upon the application of orthostatic stressors such as an active (i.e. self-induced) stand or a passive (tilt table) standing test. Lower limb muscle contractions may be important in orthostatic recovery via the skeletal muscle pump. In this study, adults with long COVID were assessed with a protocol that, in addition to the standard NCVI tests, incorporated simultaneous lower limb muscle monitoring for NMFI assessment. To accomplish such an investigation, a wide range of continuous non-invasive biomedical technologies were employed, including digital artery photoplethysmography for the extraction of cardiovascular signals, near-infrared spectroscopy for the extraction of regional tissue oxygenation in brain and muscle, and electromyography for assessment of timed muscle contractions in the lower limbs. With the novel technique described and exemplified in this paper, we were able to integrate signals from all instruments used in the assessment in a precisely synchronized fashion. We demonstrate that it is possible to visualize the interactions between all different physiological signals during the combined NCVI/NMFI assessment. Multiple counts of evidence were collected, which can capture the dynamics between skeletal muscle contractions and neurocardiovascular responses. The proposed multimodal data visualization can offer an overview of the functioning of the muscle pump during both supine rest and orthostatic recovery and can conduct comparison studies with signals from multiple participants at any given time in the assessment. This could help researchers and clinicians generate and test hypotheses based on the multimodal inspection of raw data, in long COVID and other clinical cohorts.

## 1. Introduction

The statistics from the World Health Organization (WHO)^1^ have been reflective of the devastating human and societal consequences that the COVID-19 pandemic has had for the past two years. Due to the swift pace in the research and development of vaccines, it has been possible to partially modify the natural course of the pandemic [1-3]. However, a prominent challenge is the emergence of the post-COVID-19 syndrome or ‘long COVID’ following the acute phase of the viral infection. With a plethora of debilitating symptoms across many physiological systems [4], the pathophysiology of long COVID remains elusive and subject of intense research efforts [5, 6]. Adults with long COVID can be affected by orthostatic intolerance (OI) syndromes and this could be due to dysfunction of the autonomic nervous system leading to neurocardiovascular instability (NCVI) [7, 8]. In addition, long COVID can also manifest with prolonged fatigue [9], which could be linked to neuromuscular function impairment (NMFI) [10, 11].

NCVI refers to abnormal neural control of the cardiovascular system affecting blood pressure and heart rate behaviour [12]. The current clinical NCVI assessments involve non-invasive continuous monitoring of neurocardiovascular activity (such as blood pressure, heart rate, and cerebral oxygenation levels) during orthostatic challenges, which can provide clinically useful information regarding an individual’s ability to compensate and recuperate from stressors induced by orthostatic manoeuvres [13]. Typical orthostatic stressors include the active (i.e. self-induced) stand or a passive (i.e. tilt table) standing test [14]. The autonomic nervous system provides the principal means to respond to orthostatic manoeuvres, which humans actively perform repeatedly on a daily basis [15]. Upon transiting to an upright posture, gravity causes redistribution of blood (i.e. blood pooling) in the compliant distensible veins of the abdomen and lower extremities [16]. The consequent reduction in central venous pressure leads to a decline in venous return, stroke volume and arterial pressure [17]. The primary mechanism in response to orthostatic stress is the baroreflex mediated vagal withdrawal and activation of the sympathetic nervous system, which lead to increased heart rate and systemic vascular resistance [18]. The other compensatory mechanism for combatting orthostatic challenge is termed the ‘skeletal muscle pump’. This involves the contraction of skeletal muscles of both lower limbs and abdomen, which help maintain venous return and, consequently, cardiac output by constricting underlying veins to accelerate blood flow back to the heart [19]. While the influences of the muscle pump on hemodynamic responses have been previously investigated [20-22], NMFI assessments are not routinely added to clinical NCVI assessments. Since both the autonomic nervous system and the skeletal muscle pump could malfunction in adults previously infected by SARS-CoV-2, we hypothesized that it would be important to simultaneously assess NCVI and NMFI in the study of long COVID.

In this investigation, we hence employed a wide range of continuous non-invasive biomedical technologies, including digital artery photoplethysmography for the extraction of cardiovascular signals, near-infrared spectroscopy (NIRS) for the extraction of regional tissue oxygenation in the brain and muscle, and electromyography (EMG) for assessment of timed muscle contractions in the lower limbs. Since one of the immediate challenges of this assessment was the data synchronization for cross-signal visualization, we developed and exemplified a novel technique to integrate signals from all instruments used in the assessment in a precisely synchronized fashion. Our aim was to visualize the interactions between all different physiological signals during the combined NCVI/NMFI assessment, to help researchers and clinicians generate and test hypotheses based on the multimodal inspection of raw data.

## 2. Materials and Methods

This methodological study was conducted within the wider context of a cross-sectional observational study on a participant cohort recruited for the TROPIC (Technology assisted solutions for the Recognition of Objective Physiological Indicators of post-Coronavirus-19 fatigue) investigation at Trinity College Dublin and St. James’s Hospital Dublin, Ireland. The cohort included participants who had been experiencing long COVID symptoms following the acute phase of SARS-CoV-2 infection.

Details on cohort inclusion and exclusion criteria are reported elsewhere^2^. The study received full ethical and regulatory approvals (details below).

### 2.1 Assessment protocol

The NCVI assessment protocol was comprised of a 3-minute lying-to-standing orthostatic test (active stand), consecutively followed (after a short participant break) by a 10-minute, unmedicated 70-degree head-up tilt test. A 10-minute supine rest period was required before both active stand and head-up tilt phases. For the assessment of NMFI, four 10-second maximum prompted muscle contractions of the thighs (‘squeezes’) were programmed halfway into rest periods and after both stands (Figure 1). Participants’ continuous cardiovascular and neuromuscular signals were recorded for a total duration of 35-40 minutes, with body sensor attachments as shown in Figure 2. Termination criteria were in place to stop the assessment prematurely if any adverse haemodynamic signs or symptoms were experienced by a participant.

**Figure 1.**
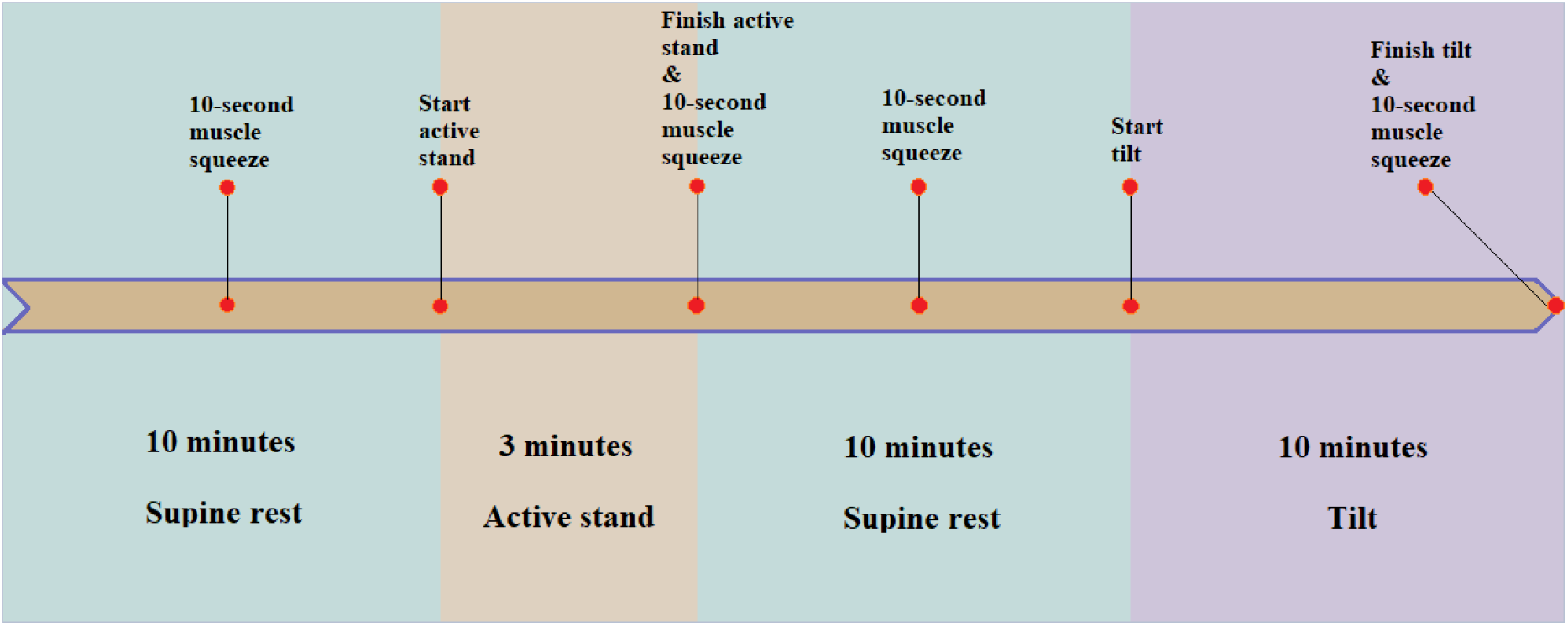
Diagram of the NCVI/NMFI assessment protocol. It was comprised of a 10-minute supine rest and a 3-minute active stand, followed by a 10-minute supine rest and a 10-minute head-up tilt. With four prompted muscle contractions of the thighs introduced at different stages of the assessment, a total of six events were included (see text for further protocol details).

**Figure 2.**
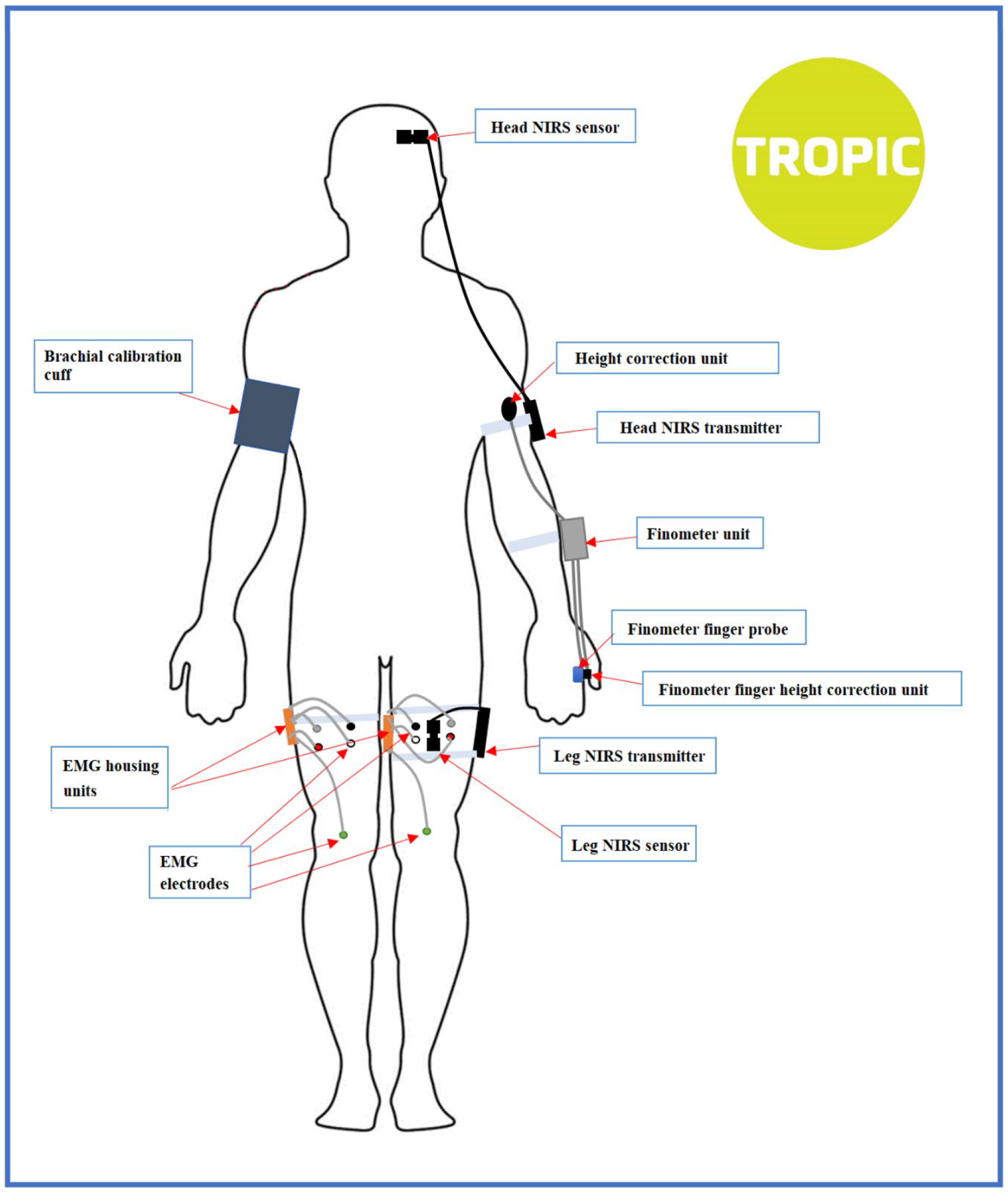
Sensor attachment diagram for the NCVI/NMFI assessment. A total of 6 modules with 15 sensors were attached to each of the participants.

#### 2.1.1 Active stand

The active stand test is a clinical procedure for assessing the spectrum of abnormal cardiovascular responses to standing and evaluating causes of orthostatic intolerance, including orthostatic hypotension [23], orthostatic hypertension [24] and postural orthostatic tachycardia syndrome [25]. The active stand test is well established as a clinical technique for the study of orthostatic intolerance in both younger and older individuals [14, 26]. For the active stand, participants underwent a lying-to-standing orthostatic test with non-invasive beat-to-beat blood pressure monitoring using digital photoplethysmography (Finapres® NOVA, Finapres Medical Systems, Amsterdam, The Netherlands). The height correction unit was zeroed and implemented as per manufacturer’s specifications. Participants underwent ten minutes of supine rest on the horizontally levelled tilt table (Agasan KT-1060/E, AGA Sanitätsartikel GmbH, Löhne, Germany) with a height of 730 mm. The first 1-4 minutes of supine rest period were also used for calibration purposes, to achieve a PhysioCal repetition rate of 70 beats or more [27]. After at least 5 minutes of uninterrupted supine rest, and a 10-second countdown, participants were asked to stand independently and as quickly as possible. The PhysioCal, which automatically calibrates the blood pressure measured at the finger cuff, was turned off just before the stand and switched back on at 1-minute post-stand. Throughout the recording, participants were asked to remain motionless and in silence with the monitored arm (left) resting extended by the side, except for reporting any symptoms of concern.

#### 2.1.2 Head-up tilt

A head-up tilt test is a simple, non-invasive examination that can be useful for the differential diagnosis of syncope and orthostatic intolerance syndromes [28, 29]. In the head-up tilt test, unlike the active stand test, the motion of the tilt table provides the means of orthostatic challenge, with little muscle activation taking place. For the tilt procedure, which was medically supervised and started after a brief (< 5 minutes) break following active stand, participants were affixed to the electrically motorised tilt table with footboard support. Approximately 10 seconds of travel time took place from 0-70°. The tilt table equipment was the same as for the active stand test (Agasan KT-1060/E, AGA Sanitätsartikel GmbH, Löhne, Germany), and powered by a single-speed electric motor controlled by a pneumatic foot switch. During the tilt, participants were secured with two straps, fastened at the levels of the knees and chest. Throughout the tilt, participants underwent Finapres® NOVA monitoring (with PhysioCal on and continuous ECG monitoring) during an initial period of uninterrupted supine rest of at least 5 minutes and a subsequent head-up tilt to 70° for 10 minutes or until symptoms developed.

### 2.2 Instrumentation

As well as the continuous non-invasive digital artery photoplethysmography for the extraction of cardiovascular signals (Finapres® NOVA, Finapres Medical Systems, Amsterdam, The Netherlands), we also used near-infrared spectroscopy (NIRS) monitoring for the extraction of regional tissue oxygenation in brain and muscle, and surface electromyography (EMG) to measure the muscle activation of the thigh muscles, both non-prompted and prompted (the latter in the form of thigh muscle squeezes that were specifically programmed into the assessment protocol). With a total of 6 modules and 15 sensors across Finometer, NIRS and EMG, the attachment locations of the sensors and the placement of the transmitters (shown in Figure 2) were kept consistent for all participants by strict application of a Standard Operating Procedure, on which all researchers were trained.

#### 2.2.1 Finometer

The Finapres® NOVA (Finapres Medical Systems BV, The Netherlands), was used in this study to measure the finger blood pressure noninvasively on a beat-to-beat basis. The Finapres® NOVA measures brachial blood pressure on command, from which corrections to the finger blood pressure are made (these oscillometric blood pressure measurements took place on the right arm during the calibration phases). The device also corrects for the hydrostatic height of the finger with respect to the heart level. The beat-to-beat data collected were extracted with the NOVAScope® v1.9 software.

#### 2.2.2 NIRS

Near-infrared spectroscopy (NIRS) provides non-invasive and non-ionizing technology that has been used to measure variations in oxygenated and deoxygenated haemoglobin concentrations in various human tissues [30-32]. It has been shown that NIRS provides consistent readings with other measurement modalities in various applications, including cerebral blood flow [33] and skeletal muscle contractions [34]. With the time-resolved, frequency-domain, and continuous wave spectroscopic implementations of NIRS that have been developed in recent years, each with its own advantages and targeted applications [35], NIRS has a potential to be widely used in both research and clinical settings due to its mobility and high temporal resolution [36].

NIRS measurements record absorbance of light at several wavelengths, where changes in absorbance in the region near 850 nm are ascribed to oxygenated haemoglobin (O_2_Hb), and absorbance in the region near 760 nm is attributed to deoxygenated haemoglobin (HHb) [37]. Combinations of the O_2_Hb and HHb are often reported to describe physiological events, including oxygen saturation (O_2_Hb/(O_2_Hb + HHb)) [38], haemoglobin difference (O_2_Hb – HHb) [39] and tissue saturation index (TSI) [40].

Many commercial and custom-built NIRS instruments may be found in the market. These systems vary in terms of their application and system engineering, with trade-offs between light sources, detectors, and instrument electronics. In our study, we used the PortaLite® (Artinis Medical Systems,

The Netherlands), which is a wireless NIRS device capable of transmitting testing data through Bluetooth at a maximum sampling frequency of 50Hz. Oxysoft v3.2.70 was used as a user’s interface for the setup, recording and exporting of NIRS data. The sampling frequency was set at 50Hz for all participants. Two identical PortaLite® devices were used in the assessment setup, as shown in Figure 2, for measuring the oxygenation of the brain (left frontal lobe) and the left thigh (biceps femoris longus), respectively.

#### 2.2.3 Surface Electromyography

Electromyography (EMG) is a form of electrodiagnostic testing that measures muscle response or electrical activity in response to a nerve’s stimulation of the muscle [41]. An electrodiagnostic signal examines the electrical currents generated in muscles during their contractions, which are interpreted as neuromuscular activity [42, 43]. The invasive intramuscular EMG, which uses needles or fine wires as a primary means for signal acquisition, is traditionally prevalent in clinical settings [44]. Surface Electromyography (sEMG) is, on the other hand, a non-invasive technique for detecting, recording, and interpreting the electrical activity of groups of muscles at rest and during action [45-47]. In its simplest sense, sEMG is a highly sensitive voltmeter that detects depolarisations and hyperpolarisations occurring on the sarcolemma, which are necessary for, and precede, the contraction of a muscle [48]. The usage of sEMG is primarily applied to research environments, with few applications found in clinical laboratories, despite its clear potential as a non-invasive measure of muscle activity [49]. It is often considered more complex to analyse than intramuscular EMG as parameters of direct clinical relevance cannot be readily extracted from the recorded signal [50].

The SHIMMER® (Shimmer Sensing, Dublin, Ireland) is a wearable wireless sensing device that can be tailored to various research and industrial applications, including EMG, ECG and accelerometer. In the setting for this research, two SHIMMER® devices were used to monitor and record EMG signals for the rectus femoris and vastus lateralis in both legs. The sampling frequency was set to 1024Hz and kept consistent to all participants. Just before starting the uninterrupted 5-minute rest phase in both active stand and tilt, and after active stand and tilt, participants were asked to perform a ten-second maximum bilateral quadriceps contraction with continuous EMG and muscle NIRS monitoring.

### 2.3 Data collection

All data were collected immediately after each assessment. NOVA, NIRS and EMG data were recorded and saved in time series format on three separate research computers. The beat-to-beat Finapres® NOVA signals were recorded in NOVAScope® v1.9 and exported in .csv format. The NOVAScope® output consists of separate data frames, including the “Basic Nova” (e.g. systolic blood pressure (SBP), diastolic blood pressure (DBP), and heart rate (HR)) and “Advanced hemodynamics” (e.g., Cardiac Output CO, etc.) files. Each data frame contains its own “Time” variable corresponding to the time lapse from the beginning of the recording. The exported NIRS data frame contains signals from the two PortaLite® devices and a sample number variable, from which the time variable was derived with a sampling frequency of 50Hz. The two EMG devices export their data files separately, with each data frame containing its own time variable.

### 2.4 Data processing

The data from NOVA, NIRS and EMG were saved *in situ* in separate research computers at the end of each recording. Without any pre-processing, these raw data were transferred securely to a local research server where data processing and visualization took place.

#### 2.4.1 Data merging

All the raw data collected was processed in R version 4.0.5 using RStudio 1.4.1106 (Boston, MA). The entire dataset consisted of three main groups of time series data files, namely the NOVA, NIRS and EMG. As a convention adopted for this study, the data merging process began with the NOVA data files, where data frames including “Basic Nova” and “Advanced hemodynamics” were merged into one data frame by the time variable associated with each data frame. Similarly, the EMG data recorded for the left and right thighs were merged into one data frame. With NOVA, NIRS and EMG data neatly organized, all three data frames were merged into one by the “Time” variable present in each data frame, forming the full data frame containing 70 variables with over 2 million observations for each participant’s assessment. A consistent naming convention was adopted for the columns during merging, which was particularly helpful in streamlining the data syncing process.

#### 2.4.2 Three-stage data syncing

Although the newly merged full data frame contained all the variables from the assessment, it could not be used straightaway for visualization without precise syncing procedures across the entire data frame. Hence, a bespoke data syncing procedure was developed that consisted of three processes: instrument-level data syncing, fully merged data syncing, and cross-group data syncing (Figure 3). As the diagram in Figure 4 shows, a timer was used for syncing signals between NOVA, NIRS and EMG, by manually pushing a key on the keyboard of each computer upon a 5-second countdown approaching each event during the assessment.

**Figure 3.**
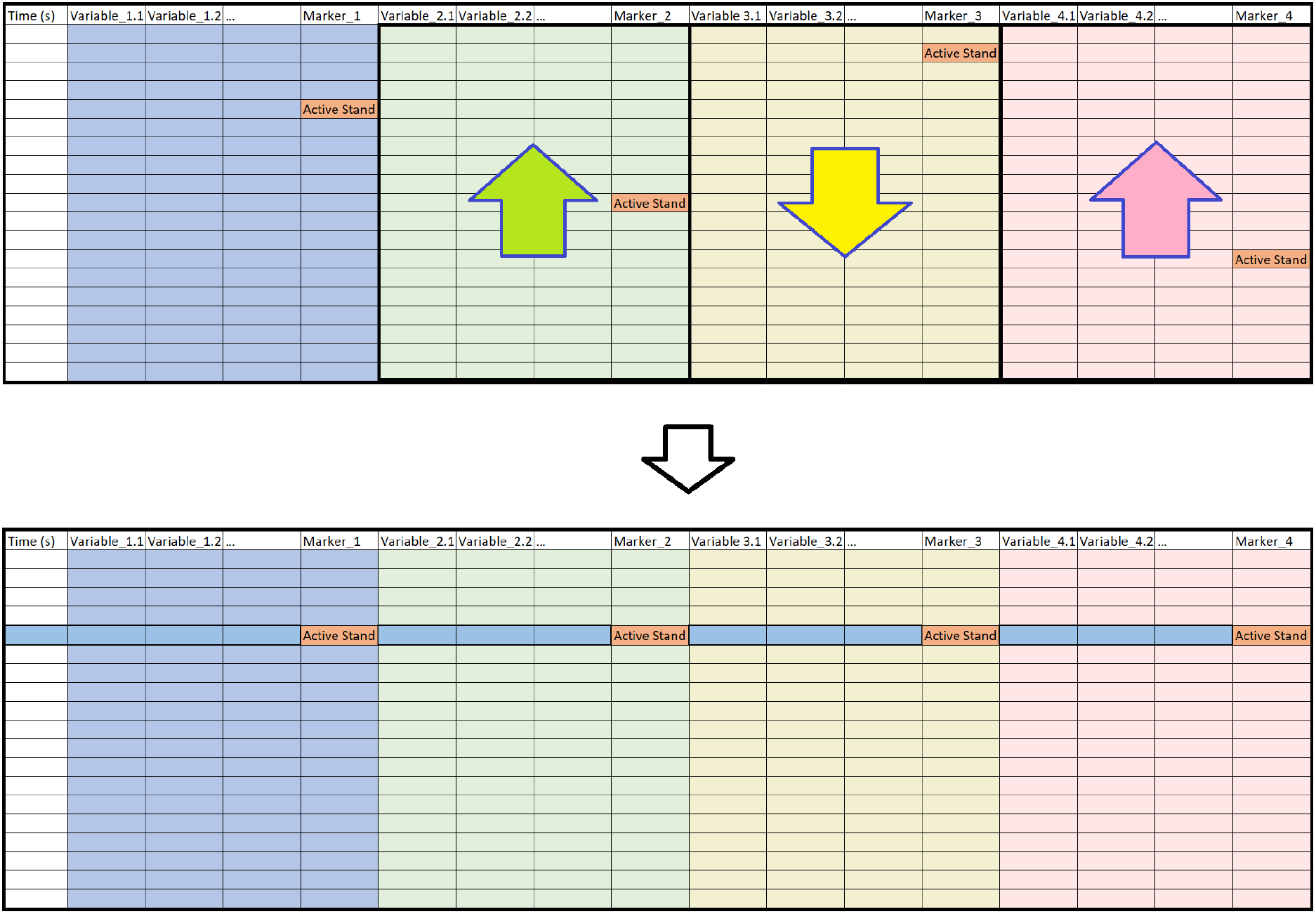
Illustration of the data syncing technique used in this study. A data frame that is merged from four separate time series data frames is shown on the top. There is an event marker variable “Active Stand” in each of the four constituent data frames, the time of which is located at different observations. The data frame is synced by shifting the variables in each of constituent data frames, so that all the event marker variables are aligned, as shown at the bottom.

**Figure 4.**
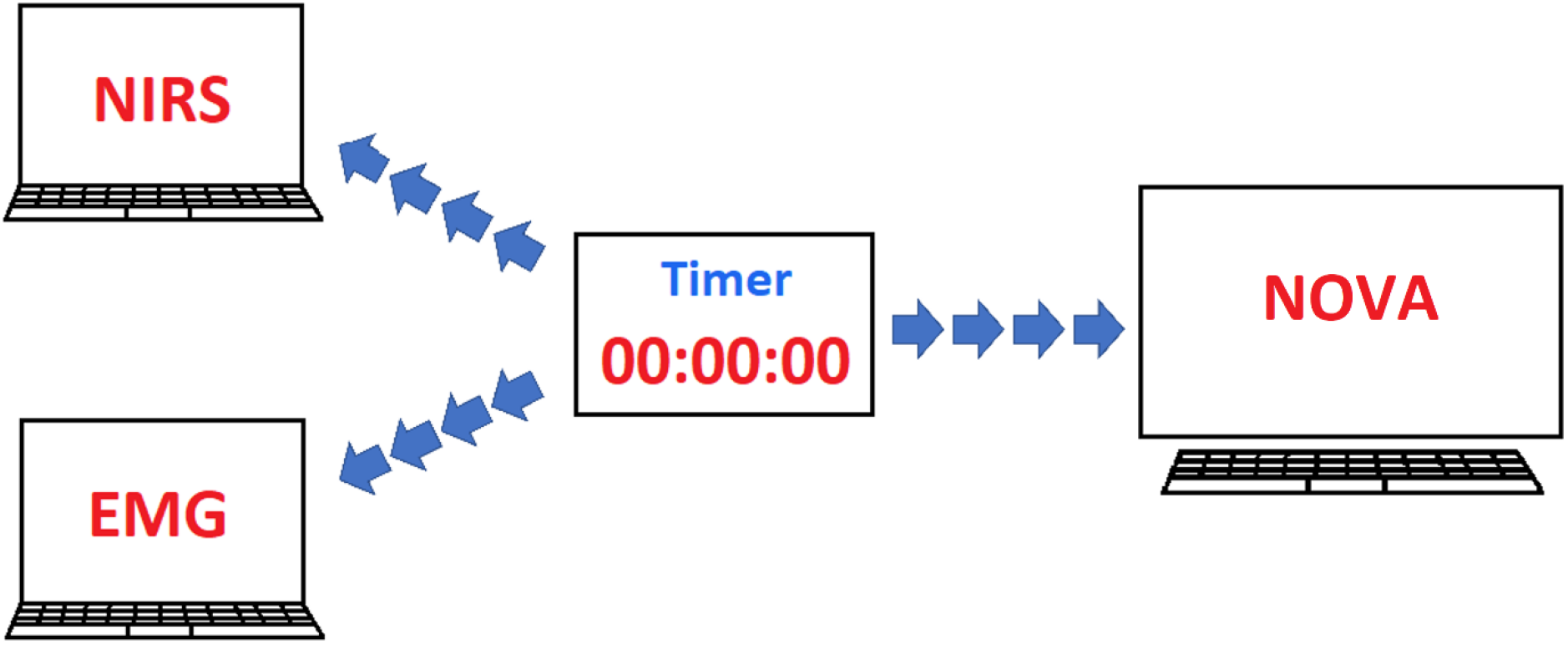
Communication diagram between different devices in the NCVI/NMFI setup. A timer was used for syncing signals between NOVA, NIRS and EMG, by manually pushing a key on the keyboard of each computer upon a 5-second countdown approaching each event during the assessment.

##### 2.4.2.1 Instrument-level data syncing

As a first step, the NOVA data frame was synced. This was done by shifting the columns originated from each data file, so that all event markers appearing in the merged data frame were aligned, as illustrated in Figure 3 where, for illustration purposes, the event “Active Stand” is used for aligning the data frames. However, in practice, syncing could be done by any other event or moment in the assessment protocol. It is by the same method that the merged EMG data frames were synced.

##### 2.4.2.2 Full data frame syncing

With the device-level data synced, the columns originated from NIRS and EMG data frames were adjusted, so that the “Active Stand” event markers across all constituent data were aligned within the final data frame. Due to the difference in sampling frequencies between NOVA, NIRS and EMG signals, the final data frame contained missing values, which is of relevance for the subsequent visualization.

##### 2.4.2.3 Cross-group data syncing

After obtaining a full precisely synced data frame for each participant, it was then possible to perform cross-group comparison studies. Using the same technique, fully synced data frames of different participants can be merged and synced at any given time. Since it is often the case that the merged data frame is quite large at this stage, a good naming convention was key to organise the variables. If the variables of interest are already known, creating subsets of the full data frames prior to merging is advised.

### 2.5 Visualization for comparative studies

Aiming to demonstrate the practicality of the proposed methodology for comparative studies, we arbitrarily selected assessment data from two participants, including a participant feeling fatigued and a participant who did not report fatigue as part of their long COVID symptomatology. It should be noted that the purpose of the comparison was by no means to extract clinical conclusions on the effect of fatigue on physiological responses to the orthostatic challenges, but to exemplify the proposed data syncing and visualization technique.

## 3. Results

### 3.1 Assessment visualization

With the bespoke data merging technique detailed above, a general overview of physiological behaviours during the NCVI/NMFI assessment can be efficiently visualized within the same plot, as exemplified in Figure 5 for the assessment of a participant who had been experiencing periodic postural orthostatic tachycardia symptoms. In this case, we chose to visualize (from top to bottom) EMG (left leg: channels 1 and 2), TSI of left leg and left frontal lobe, and SBP, DBP, CO and HR. With cross-station signals fully integrated and precisely synced, possible interactions between different physiological signals in response to orthostatic stresses can be evaluated. Thanks to the addition of EMG signals, we were able to evidence the dynamics between muscle contractions of the thigh (both prompted and non-prompted) and neurocardiovascular responses throughout the assessment.

**Figure 5.**
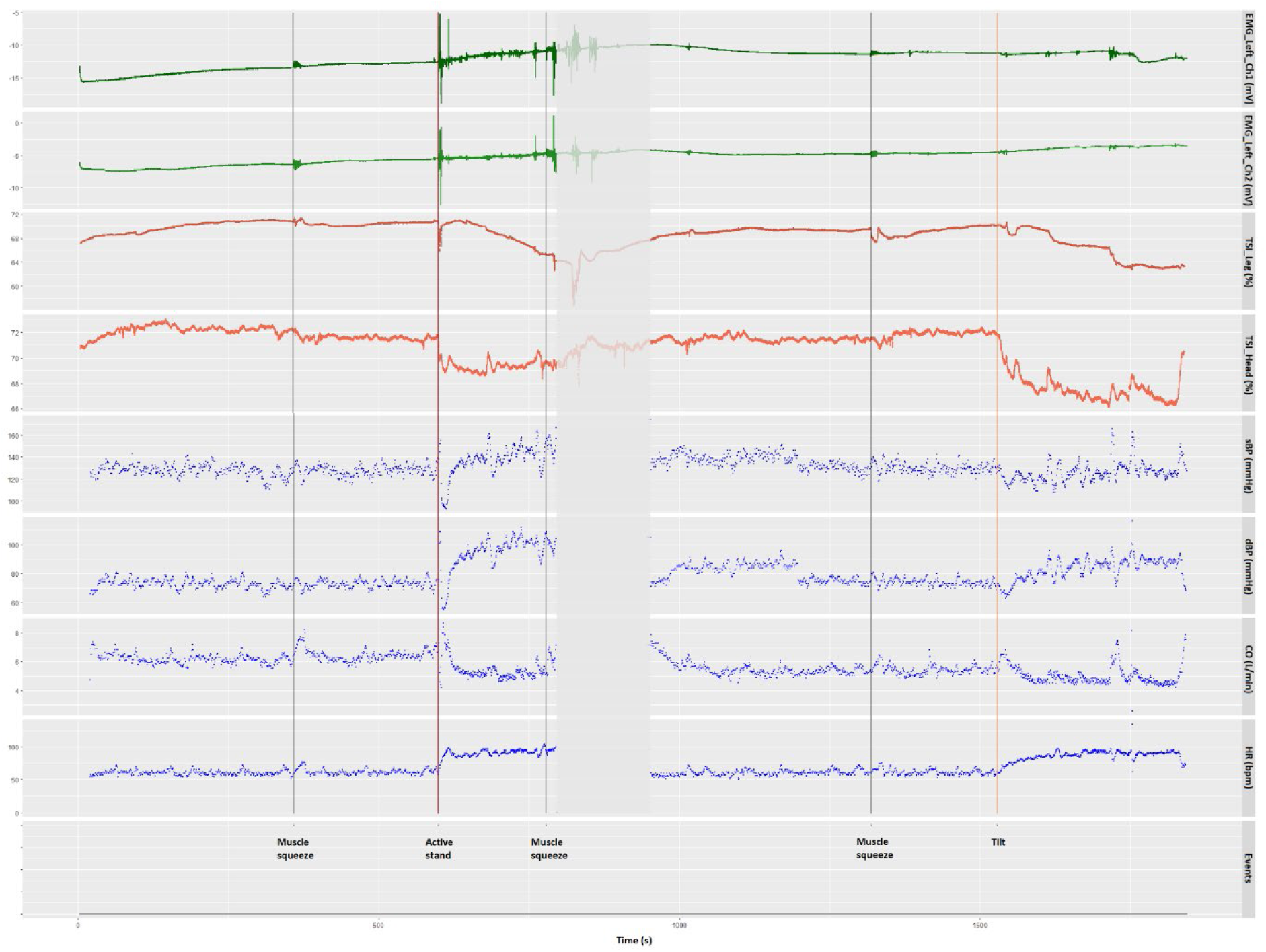
Visualization of integrated signals recorded from different instruments used in the NCVI/NMFI assessment, including the EMG amplitude of the left vastus lateralis (EMG_Left_Ch1) and rectus femoris (EMG_Left_Ch2), the TSI of the left thigh (TSI_Leg) and the left forehead (TSI_Head), the systolic blood pressure (SBP), the diastolic blood pressure (DBP), the cardiac output (CO) and the heart rate (HR) of a participant. The vertical lines indicate the time at which all events took place, including active stand, head-up tilt and prompted thigh muscle squeezes, annotated at the bottom of the plot.

As soon as the active stand took place (red vertical line in Figure 5), the participant experienced a transient (initial) drop in both SBP and DBP, which seemed to overshoot baseline readings afterwards. Derived cardiac output data showed the opposite trends. The heart rate increased immediately after the participant stood up before reaching a maximum of approximately 100 bpm within 20 seconds. This orthostatic tachycardia persisted after the participant recovered the baseline blood pressure levels. As indicated by the EMG green lines, muscle activation was evident during the pre-active stand “squeeze” and throughout the active standing phase, potentially reflecting changes in muscle sympathetic nerve activity [51, 52]. Post-active stand, there seemed to be a sudden reduction in left frontal lobe TSI and a gradual reduction in left leg TSI.

As indicated by the breaks in blood pressure and heart rate signals between active stand and head-up tilt (shown as the greyed-out area in Figure 5), the beat-to-beat NOVA recording was paused for approximately 3 minutes to mitigate the discomfort felt by the participant’s finger whilst leaving the finger cuff mounted and ready for the tilt phase.

Post tilt, the participant experienced a gradual increase in heart rate, which was sustained above baseline values until the test had to be prematurely terminated given ongoing symptoms of orthostatic intolerance. As the green EMG lines show, during the tilt there were a few non-prompted muscle contractions of the thigh muscles. Post-tilt there also seemed to be a sudden reduction in left frontal lobe TSI and a more gradual reduction in left leg TSI, possibly with more sudden “dips” in the latter associated with non-prompted contractions of the thigh muscles. Apparent peaks in cardiac output and SBP were also suggested at the time of muscle contractions. In terms of SBP and DBP behaviour during tilt, they both seemed to have increased variability with a possible overshoot in the latter.

### 3.2 EMG comparison

For exemplification purpose, the EMG signals of a fatigued and a non-fatigued participant were compared by syncing at the onset of the active stand test, as shown in Figure 6. Without any pre-processing, all four EMG signals shown in Figure 6 (two on each leg) contain noise and artefact of different origins. Generally, extrinsic sources that can be influenced by experimenters are responsible for most of the imperfections, including electrode selection, interelectrode distance and interfacial preparation between the electrodes and the skin [50, 53]. It is also obvious that the baseline readings of the four EMG spectra are located in different amplitude ranges, which is likely to be caused by the difference in interelectrode distances resulted from slight differences in electrode placement [54]. In addition, anatomical variances are an additional source of intrinsic noise, which may complicate inter-subject comparisons [50]. The effect of these noise and artefacts can be mitigated with different signal pre-processing techniques that are commonly applied to EMG spectra, including normalization, filtering and segmentation [55]. Despite the noise and artefacts associated with the raw signals, these time series EMG spectra shown in Figure 6 can be adequately used for detecting the time of muscle contraction [56, 57], both prompted and non-prompted, which was the original purpose of using surface EMG in this study.

**Figure 6.**
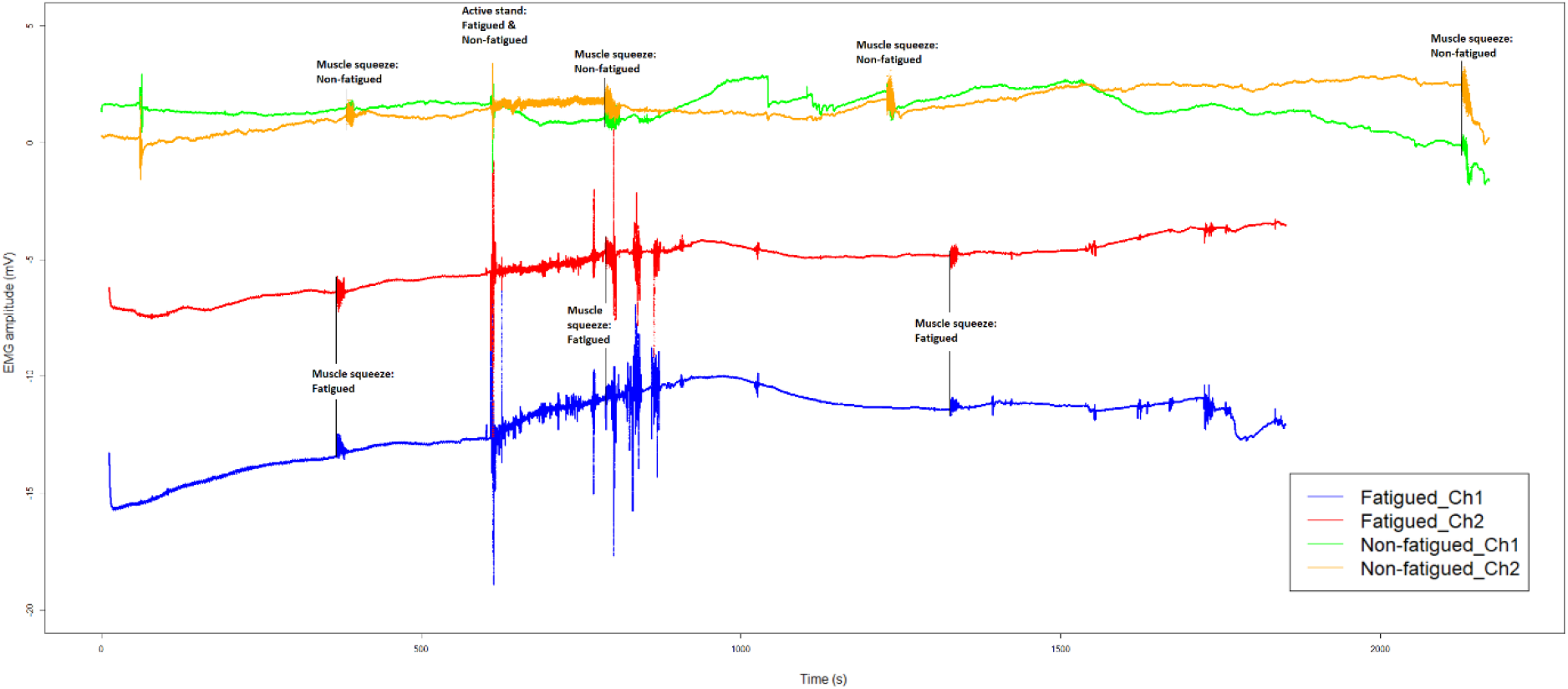
Surface EMG signals of the left vastus lateralis (shown as Ch1) and the left rectus femoris (shown as Ch2) of a fatigued participant, compared with a participant who did not report fatigue as part of the long COVID symptomatology. Thigh muscle contractions that were captured during the two recording sessions are marked by black vertical lines with corresponding annotations. The two data frames are synced at the onset of the active stand test at approximately 600 seconds.

Combining the epochs of the 10-second prompted contractions of the thigh reveals the characteristics of each active muscle squeeze performed during the assessment (see Figure 7, where the fourth contraction is not shown for the fatigued participant due to early tilt termination). Centred by a 10-second prompted muscle contraction, each epoch contains a 30-second unprocessed time series EMG recording. Although EMG signals without normalization may not be appropriate for cross-group comparisons, the amplitude of an EMG signal from a given muscle with the same electrode setup in the same recording session can be compared [58-60]. As suggested by Figure 7, the EMG amplitude in the fatigued participant seems to have a greater decline over time during the 10-second contraction, as well as showing greater signal artifact during the second squeeze. Even though the visualization in itself cannot be explanatory from a pathophysiological perspective, one may hypothesize from the visualization that the deterioration in the action potential in the fatigued participant over time could be an indication of underlying NMFI caused by a reduction in motor unit recruitment or firing rate [50].

**Figure 7.**
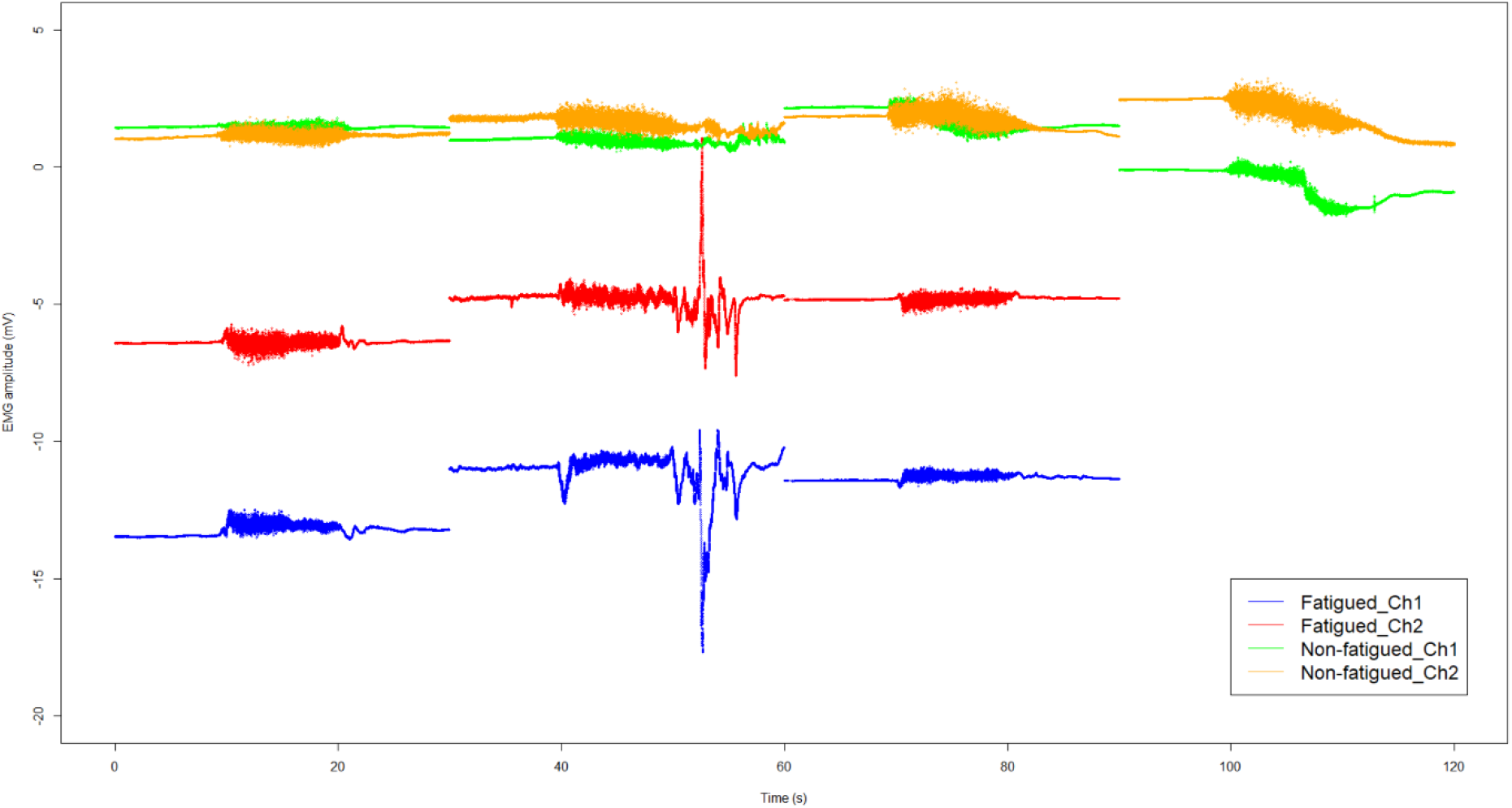
Surface EMG signals of the left vastus lateralis (shown as Ch1) and the left rectus femoris (shown as Ch2) of a fatigued participant, compared with those of a participant who did not report fatigue. An epoch of 30 seconds EMG signals is extracted for each 10-second prompted contraction of the thigh, together with 10 seconds before and after the muscle contraction. The fourth muscle squeeze for the fatigued participant was not performed due to early termination of the head-up tilt test.

## 4. Discussion

In this methodological study, we demonstrate that it is possible to visualize multiple independent physiological signals during a combined NCVI/NMFI assessment, which can capture the dynamics between skeletal muscle contractions and neurocardiovascular responses throughout the assessment. A proposed bespoke multimodal data visualization can offer an overview of the functioning of the muscle pump during both supine rest and orthostatic recovery and can conduct comparison studies with signals from multiple participants at any given time in the assessment. This could help researchers and clinicians generate and test hypotheses based on the multimodal inspection of raw data, in long COVID and other clinical cohorts.

### 4.1 Raw data visualization for NCVI/NMFI assessment

The proposed data merging and visualization technique was applied to the raw data directly exported from software packages for NOVA, NIRS and EMG recordings. Without further processing, the final data frame captures a comprehensive set of information at the highest possible definition.

It has been the norm in many fields that raw data collected from assessments are pre-processed before visualization takes place, which increases processibility in data analysis [61], enhances the visual fidelity in presenting the results [62] and reduces the cost of storage and maintenance of the database [63]. However, one of the major disadvantages of data pre-processing is that it could lead to over-processing, thereby causing undesirable data loss that may be of pathophysiological and/or clinical relevance [64, 65]. With raw data visualization, on the other hand, it is possible to integrate large data sets and visualize all information at the same time at the highest resolution and without losing any detail of the signals, thanks to the recent development of computational capabilities and software packages [66]. Indeed, our data visualization method was designed to show all relationships between signals in a summarized way. By also showing possible data artifacts, this visualization method defers to the researcher/clinician to comment on their possible physiological relevance, especially in new conditions such as long COVID where there is not much pre-existing medical literature. Indeed, we believe that a full raw data visualization approach is more appropriate than taking out or pre-filtering presumed artefactual signals from the outset, which is often done by non-clinicians [67]. In addition, by “zooming out” the raw data visualization, transient signal artifacts are easier to detect (and sometimes lost to the naked eye), providing the clinician/researcher with a further insight as to the variability of the raw physiological signals at hand.

In the context of the NCVI/NMFI assessment, where so many physiological systems are expected to dynamically interact, we believe that the risks associated with not pre-processing the signals outweighs the benefit of data pre-processing in visualising the NCVI data, at least for hypothesis generation purposes. As demonstrated, the proposed methodology is sufficient for visualizing raw data from the multimodal NCVI/NMFI assessment without any need of signal pre-processing. We can seamlessly integrate participants’ NOVA, NIRS and EMG signals in a precisely synced fashion, from which the relationship between different physiological responses could be visually examined for the purpose of generating pathophysiological hypotheses. Furthermore, our methodology also provides a platform where precisely synced assessment data can be conveyed to the next stage for data processing and bespoke analyses in specific applications, as desired. Given the fact that the interactions between muscle contraction and neurovascular physiological responses to orthostatic stressors are relatively understudied, the proposed raw data visualization methodology could pave the way for the further study of the role of the skeletal muscle pump in orthostatic intolerance syndromes and inform future clinical guidelines.

Despite the potential advantages of raw data visualization in NCVI/NMFI assessment, it requires moderately high computational assets to perform the tasks. Some coding skills in statistical software packages are also needed for processing the data. With the EMG recording at 1024 Hz, the integrated data frames are quite large and therefore requires a sizable secure storage space.

### 4.2 Data syncing

The merged data frames were synced by the events, which were manually marked by a timer with 5-second countdowns across three different instruments during each assessment. Despite every effort spent (as per Standard Operating Procedure) on the best possible synchronization of the key pushes, differences in reflex and human errors are inevitable at the operational level. While this issue could be potentially mitigated by scripting a single key push for marking events across all three sets of equipment, the integration of these systems could evoke complications in the clinical facility where the assessments took place. Hypothetically speaking, even if all three systems were integrated with seamlessly event calling commands, the signals written in the results files would not have been synced perfectly, which is the case for the NOVA data files. Despite having been validated and used in previous studies [68-70], the data files exported from the NOVA may have a discrepancy of as much as 2 seconds between different signals. This is most likely an intrinsic issue caused by the modular architecture of both the hardware and software. With each component reporting its parameters to separate files, coupled with the fact that different calculation methods are used for the derived variables, the results files are written to the designated folders at different time stamps, as confirmed by the manufacturer. As a result, manual data syncing would still be needed before any processing and analysis could be done, regardless of the precision of syncing methods. Furthermore, integrating multiple Bluetooth devices with a slave configuration would very likely cause connection instability and random packet loss [71], which would further limit the permissible sampling frequency of its constituent devices. Therefore, we believe that it was appropriate to use the proposed manual syncing method, together with the precise data syncing procedures described in the present manuscript.

### 4.3 EMG sampling frequency

Bluetooth Low Energy (BLE), one of the most popular wireless data transmitting technologies, was used for both NIRS and EMG, thanks to its high data transmission rate, low power consumption, strong signal strength, miniaturized size, and low cost [72]. Due to the nature of Bluetooth connection, communication between the wearable modules and the host computer was limited to certain values, above which an unacceptable level of data loss can occur [73, 74]. Therefore, the EMG data used in this study were extracted from the Secure Digital (SD) cards that were embedded into the SHIMMER® devices. However, due to the hardware architecture of these EMG devices, which use serial peripheral interface as the means for data communication between the microcontroller and the SD card, the characteristics of the data logging process set certain limits to the sampling frequency of the recorded analog signal to avoid excess packet drops [75]. This leads to a trade-off between sampling frequency, which dictates the details of the signals captured, and packet transmission, which governs the quality of the data received in the recording [76]. In general, high sampling frequencies tend to increase packet drops, which is a measure of data loss during the recording. It was discovered that setting the sampling frequency of EMG measurement to 512 Hz yielded a data loss of below 1%. However, it is debatable whether 512Hz could capture sufficient EMG information for physiological research [77]. It was also found that increasing the sampling frequency of the EMG recording to 2048 Hz, which is recommended by previous studies [44, 50], resulted in a packet drop of over 10%, which is below the minimum required for EMG analysis, based on the manufacturer’s manual. As a result, a sampling frequency of 1024 Hz was used for the EMG recording, which returned an acceptable data transfer rate of 96.5% according to the manufacturer’s recommendations and was consistent with those used in previous investigations [78, 79]. However, despite meeting the minimum requirement for EMG recording based on the Nyquist-Shannon Sampling Theorem, a sampling frequency of 1024 Hz is not optimal for capturing the exhaustive details of all muscle activities [44]. It is, however, appropriate for detecting muscle activation and amplitude monitoring [77], which were the sole usage of the EMG signals in this study.

## Conclusion

A novel bespoke raw data visualization methodology was presented for combined NCVI/NMFI assessments of participants experiencing long COVID symptoms. This allowed functional visualization of both the autonomic nervous system and skeletal muscle pump in response to orthostatic stresses induced by the active stand and head-up tilt tests. We illustrated that signals from various instruments used in the NCVI/NMFI assessment, including NOVA, NIRS and EMG, can be seamlessly integrated and precisely synced. With the addition of EMG signals to the traditional NCVI assessment, we collected novel evidence that may help the understanding of the dynamics between muscle contractions of the thighs (both prompted and non-prompted) and neurocardiovascular responses throughout the assessment, which can potentially provide insights into the functioning of the skeletal muscle pump both during supine rest and in the recovery phase. Avenues for further refinement include the semi-automation of this method to reduce the visualization processing time and the subsequent dissemination of the proposed data visualization technique to a wide range of applications where precise cross-platform data syncing is required.

## Data Availability

All data produced in the present study are available upon reasonable request to the authors

## Acknowledgments

This study (Technology Assisted Solutions for the Recognition of Objective Physiological Indicators of Post-Coronavirus-19 Fatigue: TROPIC Study) was funded by a Grant from Science Foundation Ireland (SFI) under Grant number 20/COV/8493 and supported by SFI Grant number 18/FRL/6188. The funder had no role in the conduct of the research and/or preparation of the article; in study design; in the collection, analysis, and interpretation of data; in writing of the report; or in the decision to submit the paper for publication. We are very grateful to our participants for their involvement in the study.

## Ethical approval

This study received full approval by the St James’s Hospital/Tallaght University Hospital Joint Research Ethics Committee (Submission Number: 104: TROPIC; Approval Date: 4 May 2021) and the St James’s Hospital Research & Innovation Office (Reference: 6566; Approval Date: 14 May 2021). The study was performed in accordance with the ethical standards laid down in the 1964 Declaration of Helsinki and its later amendments. All participants gave their informed consent prior to their inclusion in the study. All aspects of the study were executed in compliance with the General Data Protection regulation (GDPR), and Irish regulations including the Health Research Regulations and the Data Protection Act 2018.

## Author contributions

Conceptualization: Roman Romero-Ortuno, Ann Monaghan; Methodology: Roman Romero-Ortuno, Ann Monaghan, Feng Xue, Glenn Jennings; Clinical data collection: Ann Monaghan, Glenn Jennings, Feng Xue, Eoin Duggan, Roman Romero-Ortuno; Writing – original draft preparation: Feng Xue; Writing – review and editing: Ann Monaghan, Glenn Jennings, Feng Xue, Tim Foran, Eoin Duggan, Lisa Byrne; Funding acquisition: Roman Romero-Ortuno; Resources: Roman Romero-Ortuno; Supervision: Eoin Duggan, Lisa Byrne, Roman Romero-Ortuno.

## Footnotes

1 WHO Coronavirus (COVID-19) Dashboard: https://covid19.who.int/

2 https://clinicaltrials.gov/ct2/show/NCT05027724

